# Which Attention Helps? A Slide-Level Ablation of Attention-Augmented CNNs for Serous Ovarian Carcinoma Grading

**DOI:** 10.64898/2026.04.21.26351441

**Authors:** Aiswarya Rani, Shivangi Mishra

## Abstract

**Background:** Distinguishing high-grade (HGSC) from low-grade serous carcinoma (LGSC) is clinically consequential yet difficult, as the subtypes share a serous morphology but differ in prognosis and treatment. Prior deep-learning studies report near-perfect accuracy, but often under patch-level evaluation that leaks slide information between training and testing.

**Methods:** We present a leakage-controlled, slide-level ablation of attention-augmented CNNs for this binary task on the UBC-OCEAN cohort (259 whole-slide images; 217 HGSC, 42 LGSC). Under an identical protocol with five-fold slide-grouped cross-validation, Macenko stain normalisation, and slide-level pooling, we compared a from-scratch backbone with no attention, Squeeze-and-Excitation, channel-only CBAM and full CBAM, a lightweight CBAM variant, and an ImageNet-pretrained MobileNet with a differential-attention module. A pathology foundation model (Phikon) with attention-based multiple-instance aggregation was evaluated as a modern benchmark. Results are reported across three training seeds, with decision thresholds selected within folds, and compared using DeLong and McNemar tests with Holm correction and bootstrap confidence intervals.

**Results:** The pretrained model performed best (AUC 0.862 ± 0.005, MCC 0.460) and was the most stable across seeds. It significantly outranked both the Squeeze-and-Excitation model (DeLong, Holm p = 0.002) and the no-attention baseline (p = 0.019). Full CBAM also outranked Squeeze-and-Excitation (p = 0.010), but adding the spatial-attention branch to channel attention produced no detectable change (0.824 to 0.822; p = 0.50), and the lightweight model did not match the deep one once training variance was accounted for. The foundation-model benchmark reached AUC 0.834 ± 0.018, below the pretrained MobileNet. A colour-only classifier reached AUC 0.66, indicating part of the separability is attributable to stain and colour rather than morphology.

**Conclusion:** Under leakage-free evaluation, performance is more modest than commonly reported. Transfer learning and the avoidance of a weak channel-only configuration produced the clearest gains, while finer attention design, network size and a pathology-specific foundation model produced no detectable improvement. With only 42 LGSC slides the task remains underpowered for finer distinctions, and most architectural differences could not be resolved.

## 1. INTRODUCTION

Ovarian cancer ranks among the most lethal gynaecological cancers in women, with a median survival below three years in advanced disease and, by GLOBOCAN 2022 estimates, approximately 324,398 new cases and 206,839 deaths worldwide. The majority are epithelial in origin and are stratified into serous, mucinous, clear cell, and endometrioid subtypes. Within the serous group, two grades govern clinical management. High-grade serous carcinoma (HGSC) constitutes approximately three-quarters of cases and accounts for most ovarian cancer mortality through rapid proliferation and early peritoneal dissemination, and low-grade serous carcinoma (LGSC), which is comparatively indolent and associated with longer survival but exhibits marked resistance to platinum-based chemotherapy. As the two grades arise through distinct molecular pathways and mandate different therapeutic strategies, their accurate discrimination is a clinically consequential diagnostic task.(1)(2)(3)(4)(5)(6)(7)

This discrimination is challenging even for expert pathologists. HGSC and LGSC share a serous, papillary architecture and are separated principally by nuclear criteria. HGSC exhibits pronounced pleomorphism, prominent nucleoli, and a mitotic rate exceeding twelve mitoses per ten high-power fields, whereas LGSC displays uniform nuclei with only mild atypia. The intrinsic heterogeneity of HGSC, which encompasses a solid, pseudo-endometrioid, and transitional (SET) variant, further attenuates the morphological boundary between the grades. This difficulty carries over to automated systems, making the distinction that matters most for treatment also the one they resolve least reliably.(8)(9)(9)

Deep learning has been applied extensively to ovarian subtyping, frequently with reported accuracies approaching ceiling. A closer examination, however, reveals three systematic methodological limitations that qualify these results. First, the task is predominantly formulated as five-class subtyping, so aggregate performance is dominated by the prevalent and readily separable classes and does not isolate the HGSC-LGSC boundary; a high multiclass score is therefore consistent with poor discrimination of the rare serous grades. Second, many reported accuracies derive from image- or patch-level partitioning, in which tiles originating from a single slide are distributed across both training and test sets. Because such tiles share staining, scanner, and patient-specific characteristics, a model may exploit slide identity as a shortcut rather than learn disease morphology, yielding performance estimates that are optimistically biased and do not generalise under case-level evaluation. Third, prevailing pipelines seldom quantify predictive uncertainty, rarely compare attention or backbone configurations under matched conditions, and use explainability for illustration rather than validation. Even where large pretrained models have been carefully tuned, gains in balanced accuracy on this task are reported to be marginal(10), suggesting the problem lies less in model capacity than in how models are evaluated and understood.

What the field therefore needs is not a higher headline number, but evidence that can be trusted. Such evidence requires performance reported with proper statistical uncertainty, comparisons that isolate the effect of each design choice, and confirmation that a model bases its decisions on tumour tissue rather than incidental cues. These requirements matter beyond benchmarking, since a model intended to support diagnosis must be both reliable under honest evaluation and interpretable enough for a pathologist to trust before it can be considered for clinical use. Guided by these principles, the present work makes three contributions.

□ A slide-level evaluation protocol is implemented in which all tiles from a slide are assigned to the same fold, tile predictions are aggregated to a single slide-level prediction, and results are reported with bootstrap confidence intervals and significance tests.
□ An ablation of attention mechanisms is performed with the backbone, dataset, and training procedure held constant, varying only the attention block across CBAM, SE, a differential-attention module, and a pretrained backbone.
□ A shortcut analysis using a colour-only baseline tests whether the separability between grades reflects morphology rather than stain or scanner signatures; direct validation of the attention modules against expert tumour masks is identified as the primary extension of this work.

## 2. RELATED WORK

Treatment of ovarian carcinoma is increasingly guided by histological subtype and grade, which makes accurate diagnosis a prerequisite for appropriate care. That diagnosis, however, still rests on the visual assessment of haematoxylin and eosin stained tissue, and is known to be difficult to reproduce. Among pathologists without gynaecological subspecialty training, agreement on ovarian carcinoma subtype is only moderate, and the two-tier system used to separate high-from low-grade serous carcinoma has itself shown measurable inter- and intra-observer variability, since it depends on fine nuclear judgements such as atypia and mitotic activity that are inherently subjective. Because of this uncertainty, pathologists frequently rely on ancillary immunohistochemical stains and second opinions to reach a confident diagnosis, which lengthens the diagnostic pathway and adds cost. These pressures, compounded by a global shortage of specialist pathologists, have motivated the search for automated tools that can support and standardise histological assessment. Digital pathology has taken up this challenge. Convolutional networks applied to whole-slide images have moved from tumour detection and tissue segmentation to full histotype classification, and the strongest systems now approach the reproducibility of subspecialist pathologists for the five-class subtyping task, exceeding the agreement typically achieved by general pathologists even when tested on tissue prepared at a different institution. This progress establishes that automated ovarian subtyping is feasible at a clinically meaningful level.(11)(9)(10)(10)

That success, however, is largely of coarse subtyping, in which the common and morphologically distinct classes are separated. The finer distinction that lies within the serous group, between high-grade and low-grade serous carcinoma, has received far less dedicated attention despite being the boundary of greatest therapeutic consequence. It is also intrinsically the hardest, as the two grades share a serous, papillary appearance and are separated mainly by subtle nuclear detail, and as high-grade tumours are themselves heterogeneous through the SET variant. This difficulty is visible even in automated results. In the analysis accompanying the dataset used here, agreement between model prediction and reference diagnosis was lowest for low-grade serous carcinoma of all subtypes. The remainder of this section examines how deep learning has been applied to this specific distinction, and where those efforts fall short.(8)(11)(12)

A substantial body of work has applied deep learning to ovarian histopathology and reported strikingly high accuracy, while contributing genuinely useful methodological ideas. Table 1 summarises the representative approaches.

**Table 1.** Representative deep-learning studies on ovarian and whole-slide histopathology, grouped by evaluation protocol.

| Study | Task | Approach | Data and evaluation | Reported result | Relevance to this work | Ref. |
|---|---|---|---|---|---|---|
| A. High reported accuracy, image-level evaluation |  |  |  |  |  |  |
| Behera et al. | 5-class subtyping (UBC-OCEAN) | EfficientNet-B0 features + fine KNN classifier | 725 images; single 80/20 split; no cross-validation | 100% accuracy; HGSC test sensitivity 42.9%, HGSC AUC 0.69 | Aggregate accuracy hides weak per-class results | (27) |
| OvCan-FIND | Subtyping | EfficientNetV2 with SE blocks | 5,942 images, 42 patients; image-level split | 99.74% accuracy | Very small test partition; authors list overfitting risk and no uncertainty estimation | (15) |
| Abd El-Latif et al. | Cancer vs normal | ResNet-50 + DT-RFE + fuzzy deep classifier | Single 80/20 split | 98.99% accuracy | Detection task, not subtype or grade | (16) |
| OVANet | Subtyping | Dual spatial and channel attention CNN | 508 original images augmented to about 13,000 | 99.01% accuracy and AUC | Attention for class imbalance; small original image set | (18) |
| B. Slide-level evaluation |  |  |  |  |  |  |
| Stochastic-attention MIL | 5-class subtyping (UBC-OCEAN, 538 WSIs) | Attention MIL on foundation-model features, regularised attention | 10-fold CV; TCGA-OV molecular subtyping as a second task | Balanced accuracy 0.854, κ 0.791 | Addresses attention concentration on few patches | (10) |
| Four-hospital study | Epithelial subtype diagnosis (319 slides, 152 patients) | UNI features + attention MIL; slide and patient aggregation | 5-fold CV; external test on TCGA-OV HGSC | Slide AUC 0.990, accuracy 0.918; external slide accuracy 0.794 | External cohort is HGSC only | (17) |
| Comparative evaluation | 5-class WSI subtyping | Feature extractors × aggregators × classification hierarchies | Matched comparison | Extractor choice outweighed aggregator; explicit HGSC stage helped | Supports controlled ablation | (10) |
| C. Attention, aggregation and reliability |  |  |  |  |  |  |
| OCCNet | Subtyping, imbalanced multi-centre WSIs | Ensemble attention CNN | Multi-centre cohort | Lower performance on rarest subtypes | Attention alone does not solve rare classes | (19) |
| MS-DA-MIL | WSI subtyping | Multi-scale domain-adversarial MIL, instance attention | H&E WSIs | Improved over single-scale MIL | Multi-resolution aggregation | (20) |
| HAG-MIL | WSI classification | Hierarchical attention-guided MIL across resolutions | Camelyon16, TCGA | Improved accuracy and interpretability | Multi-resolution attention | (21) |
| HipoMap | WSI classification | Slide-level representation weighting discriminative regions | WSIs | Improved accuracy and interpretability | Slide-level representation | (22) |
| MHAttnSurv | Survival prediction | Multi-head attention MIL | WSIs | Improved prognosis prediction | Attention on distinct morphology | (23) |
| ViT / KAT | WSI classification | Hierarchical self-attention; kernel cross-attention | Gigapixel WSIs | Scalable, efficient context modelling | Transformer-based attention | (24) |
| MixPatch | WSI classification | Patch mixing with proportion-based soft labels | Histopathology WSIs | Improved calibration and robustness | Calibration as a clinical requirement | (26) |

Behera et al. proposed a hybrid two-stage pipeline in which deep features from EfficientNet-B0 are classified by a fine-tuned k-nearest-neighbour model rather than a softmax layer, a design that cleanly separates representation learning from classification, and reported 100 percent accuracy across the five UBC-OCEAN subtypes. Their own per-class test results, however, are considerably weaker than this headline, with sensitivity falling to 42.9 percent for high-grade serous and 64.3 percent for endometrioid carcinoma, with a test AUC for high-grade serous of only 0.69, indicating that the aggregate figure does not reflect performance on the individual classes. The evaluation also used a single 80-to-20 split without cross-validation, on classes containing as few as forty original images before augmentation. The OvCan-FIND model contributed a task-specific architecture built on EfficientNetV2 with squeeze-and-excitation blocks, explicitly aimed at reducing overfitting, and reported 99.74 percent accuracy against a broad panel of standard backbones. Here the evaluation rests on 5942 images from 42 patients under an image-level split, leaving a test partition of only a handful of images, and the authors themselves list dataset bias, class imbalance, overfitting risk, absence of explainability, and lack of uncertainty estimation among the study’s limitations. Abd El-Latif et al. combined ResNet-50 features with decision-tree recursive feature elimination and a fuzzy deep-learning classifier, an interesting integration of fuzzy logic with deep representations, and reported 98.99 percent accuracy; this result, however, concerns the comparatively easier cancer-versus-normal detection task rather than subtype classification, and again uses a single 80-to-20 split. Together these studies advance hybrid classification, data-efficient architectures, and fuzzy-logic fusion, and confirm that deep networks can extract discriminative morphological features from ovarian tissue.(13)(30)(15)(16)

The common thread is that these accuracies are obtained on small datasets, under image-level splits without case-level grouping or cross-validation, and without reported uncertainty, conditions under which a model can score highly while learning slide-specific rather than disease-specific cues. The effect becomes apparent when evaluation is tightened. A recent model built on pathology foundation-model features, regularising its attention and validated under ten-fold cross-validation, reported a balanced accuracy of 0.854 and a Cohen’s kappa of 0.791, with a lower balanced accuracy of 0.704 when transferred to molecular subtyping on TCGA. In a separate multi-institutional study, an attention-based multiple-instance model aggregated patch predictions to the slide and patient level across four hospitals and was externally validated on high-grade serous cases from TCGA, again reaching robust rather than near-perfect performance. Under these stricter protocols, performance falls below the near-perfect figures reported elsewhere, most clearly on external data, where slide-level accuracy dropped to 0.794, indicating that the difficulty of ovarian subtyping, and of the serous grade distinction in particular, has frequently been understated. Establishing reliable performance therefore calls for leakage-controlled, slide-level evaluation with quantified uncertainty, which the present work adopts.(10)(17)

Beyond the choice of classifier, a large part of the methodological effort in this area concerns how attention and instance aggregation are used, since whole-slide images are far too large to classify directly and only a fraction of any slide is diagnostic. Attention modules address the first problem by reweighting features toward informative regions. Channel and spatial attention, exemplified by Squeeze-and-Excitation and CBAM, recalibrate feature responses to emphasise subtle morphology, and this idea has been extended specifically for ovarian subtyping. OVANet couples spatial and channel attention to mitigate class imbalance, while OCCNet introduces an ensemble attention mechanism aimed at imbalanced, multi-centre cohorts. These models confirm that attention improves discrimination, yet OCCNet’s own analysis notes that its performance still falls on the subtypes with fewest samples, underlining that attention alone does not resolve the scarcity of rare classes such as low-grade serous carcinoma.(18)(19)

The second problem, aggregating many patch-level signals into a single slide decision, is typically handled through multiple-instance learning. Frameworks such as MS-DA-MIL and HAG-MIL pool instance-level attention across multiple resolutions to preserve spatial context, while slide-level representations such as HipoMap weight discriminative regions hierarchically and attention-based survival models such as MHAttnSurv focus on morphologically distinct areas for prognosis. Transformer architectures extend the same principle through self-attention. Vision transformers apply hierarchical attention to gigapixel slides, and kernel-based variants such as KAT capture localised context efficiently. Reliability-oriented work such as MixPatch further shows that calibrated probabilities, not accuracy alone, are essential for clinical use.(20)(21)(22)(23)(24)(25)(26)

Two observations follow from this body of work and directly motivate the present study. First, although many attention and aggregation designs are individually effective, they are rarely compared on equal terms; a recent systematic evaluation on this very task found that the choice of feature extractor accounted for far more performance variance than the aggregation strategy, and that dedicating an explicit binary stage to high-grade serous detection outperformed a flat five-class formulation. This indicates that the contribution of any single component can only be established by holding all others fixed, which is the logic of a controlled ablation. Second, the faithfulness of attention is increasingly in question. Work on this dataset has raised the concern that attention may highlight shortcut regions rather than tumour, and comparative studies have begun assessing attention maps against pathologist annotations directly. Together these observations argue for evaluating attention mechanisms under a matched protocol and validating their explanations against expert ground truth, rather than assuming either their relative value or their faithfulness, which is the approach adopted here.(10)(10)

In summary, three gaps persist across the literature. The clinically decisive distinction between high- and low-grade serous carcinoma is seldom studied in isolation, and is instead absorbed into a multiclass average dominated by the common high-grade class. Reported performance is frequently obtained under image- or patch-level evaluation without case-level grouping, cross-validation, or quantified uncertainty, so headline accuracies are difficult to trust and often far exceed those of rigorously validated work. And attention mechanisms are typically adopted in isolation rather than compared under matched conditions, with their explanations shown qualitatively rather than validated against ground truth. The present study addresses these gaps together by isolating the binary high-versus low-grade task, evaluating strictly at the slide level under leakage-controlled cross-validation with reported confidence intervals and significance testing, comparing attention mechanisms while holding all other factors fixed, and validating attention against expert tumour masks.

## 3. METHODS

### 3.1 Dataset and Tile Extraction

The data were obtained from the UBC-OCEAN challenge (AIM Lab, University of British Columbia), a multi-institutional collection of ovarian carcinoma whole-slide images (WSIs) from more than twenty centres. Only the publicly released training set was used, as labels for the challenge test set are withheld. We retained only the two serous subtypes, high-grade (HGSC) and low-grade serous carcinoma (LGSC), framing the task as binary grade discrimination. To hold magnification constant, we used only WSIs (scanned at 20x) and excluded the tissue-microarray cores digitised at 40x. The final cohort comprised 259 slides: 217 HGSC and 42 LGSC. The scarcity of LGSC reflects its clinical rarity and is the principal factor bounding the statistical precision of our results.

Because a WSI spans tens of thousands of pixels per dimension, each slide was decomposed into non-overlapping 512×512 tiles at native resolution and resized to 224×224 for model input, with slides read in sequential horizontal strips to avoid loading gigapixel images in full. Each tile inherited the grade label of its parent slide. Tile retention then followed a two-branch strategy conditioned on annotation availability. For slides accompanied by the challenge’s supplemental tumour masks, a tile was kept only when its mask region was dominated by annotated tumour, yielding confirmed malignant tissue. For slides without masks, a saturation-based tissue detector discarded background, pen marking, and non-stained area, with tumour localisation instead deferred to the model’s attention mechanism. A fixed cap of 60 tiles per slide was applied uniformly across both classes to prevent any single slide, particularly the few LGSC cases, from dominating through near-duplicate tiles.

This procedure produced 15,532 tiles (13,012 HGSC and 2,520 LGSC), of which 4,260 (27.4 percent) were drawn from mask-confirmed tumour regions and 11,272 from tissue-detected regions. Every tile was tagged with its source-slide identifier, the provenance record underpinning the partitioning described below. The resulting class ratio was inherited directly from the slide-level distribution and deliberately not rebalanced through tile duplication.

**Fig 3.1.**
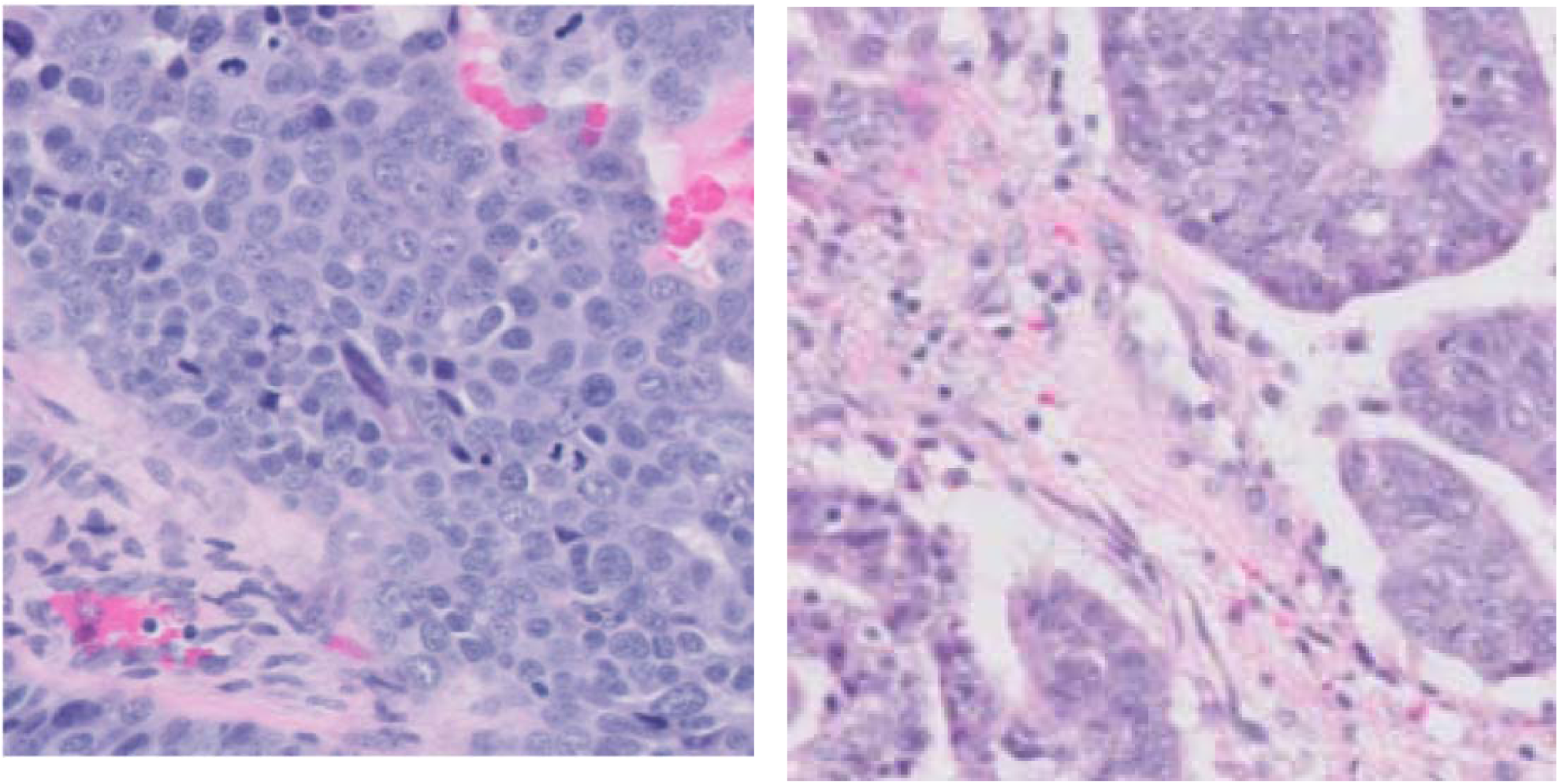
Representative H&E-stained tiles of (a) high-grade serous carcinoma, showing pleomorphic nuclei with prominent atypia, and (b) low-grade serous carcinoma, showing more uniform nuclei.

### 3.2 Leakage-Controlled Partitioning

A central priority of this study was the elimination of information leakage between training and evaluation. Because all tiles from one slide share stain, scanner, and patient characteristics, partitioning tiles at random allows a model to recognise slide identity rather than disease morphology, inflating apparent performance. To preclude this, all partitioning was performed at the slide level, so that every tile of a given slide was assigned in full to a single partition. As each WSI in this cohort corresponds to a distinct case, slide-level grouping is functionally equivalent to patient-level separation.

Evaluation used five-fold stratified group cross-validation, with the slide identifier as the grouping key and grade as the stratification target. This guaranteed that no slide contributed tiles to more than one fold while keeping the HGSC:LGSC ratio comparable across folds, with per-fold test composition ranging from six to ten LGSC slides. Within each fold, a further slide-disjoint split reserved a portion of training slides for validation, used for early stopping and learning-rate scheduling. Disjointness of the training, validation, and test slide sets was verified programmatically for every fold.

**Fig 3.2.**
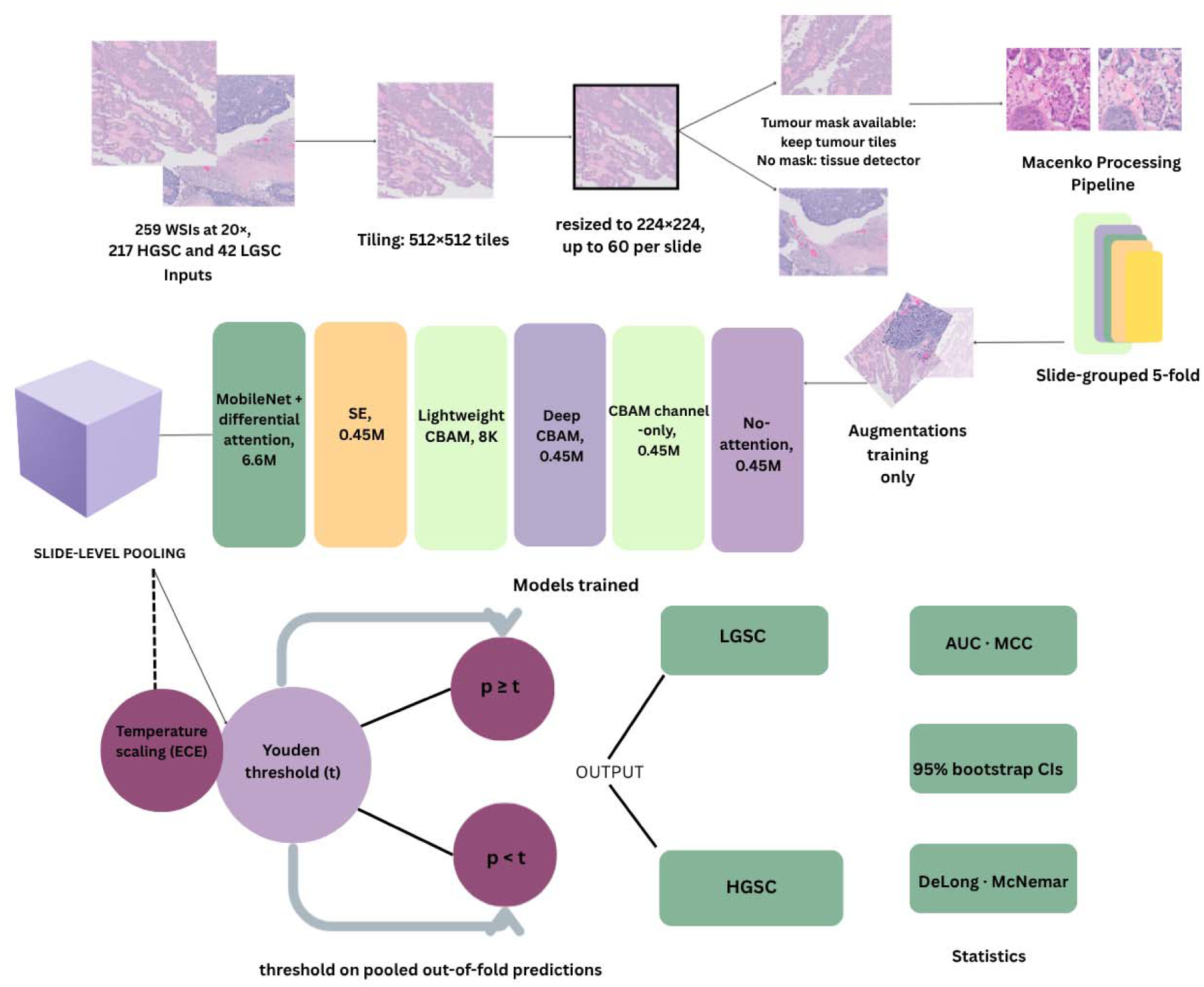
Study pipeline. Whole-slide images were tiled using mask-guided or tissue-based selection, stain-normalised, and partitioned into five slide-grouped folds. Seven models were trained under an identical protocol across three seeds; tile predictions were pooled to the slide level and thresholded within folds by Youden’s J statistic. Models were compared using bootstrap confidence intervals and DeLong and McNemar tests with Holm correction, followed by post-hoc temperature scaling. A colour-only baseline was used as a shortcut control.(28)

### 3.3 Stain Normalisation and Augmentation

Given the substantial stain variability of this multi-centre cohort, and the presence of markedly under-stained slides, Macenko stain normalisation was applied, decomposing each tile in optical-density space and re-expressing it against a reference stain matrix. Optical density was computed as;(29)

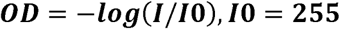

with I the pixel intensity and I0 = 255 for 8-bit images, and the dominant hematoxylin and eosin directions were recovered by singular value decomposition of the optical-density matrix,

Normalisation was performed once, before cross-validation, with every tile mapped to the stain matrix of a single well-stained tumour tile selected in advance, and the normalised tiles were stored for all subsequent experiments. Because the reference contributes only a stain matrix and carries no label information, sharing it across folds does not introduce label leakage. As the study is restricted to 20x images, a single parameterisation sufficed.

Augmentation was applied on the fly to training tiles only and comprised horizontal and vertical flips, ninety-degree rotations, and mild brightness and contrast perturbation, reflecting the rotational and staining invariances intrinsic to histopathology:

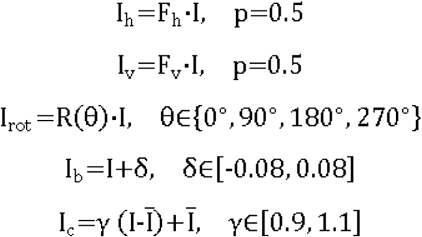

Validation and test tiles received no augmentation. A fixed random seed of 42 governed all stochastic operations for reproducibility.

**Fig 3.3.**
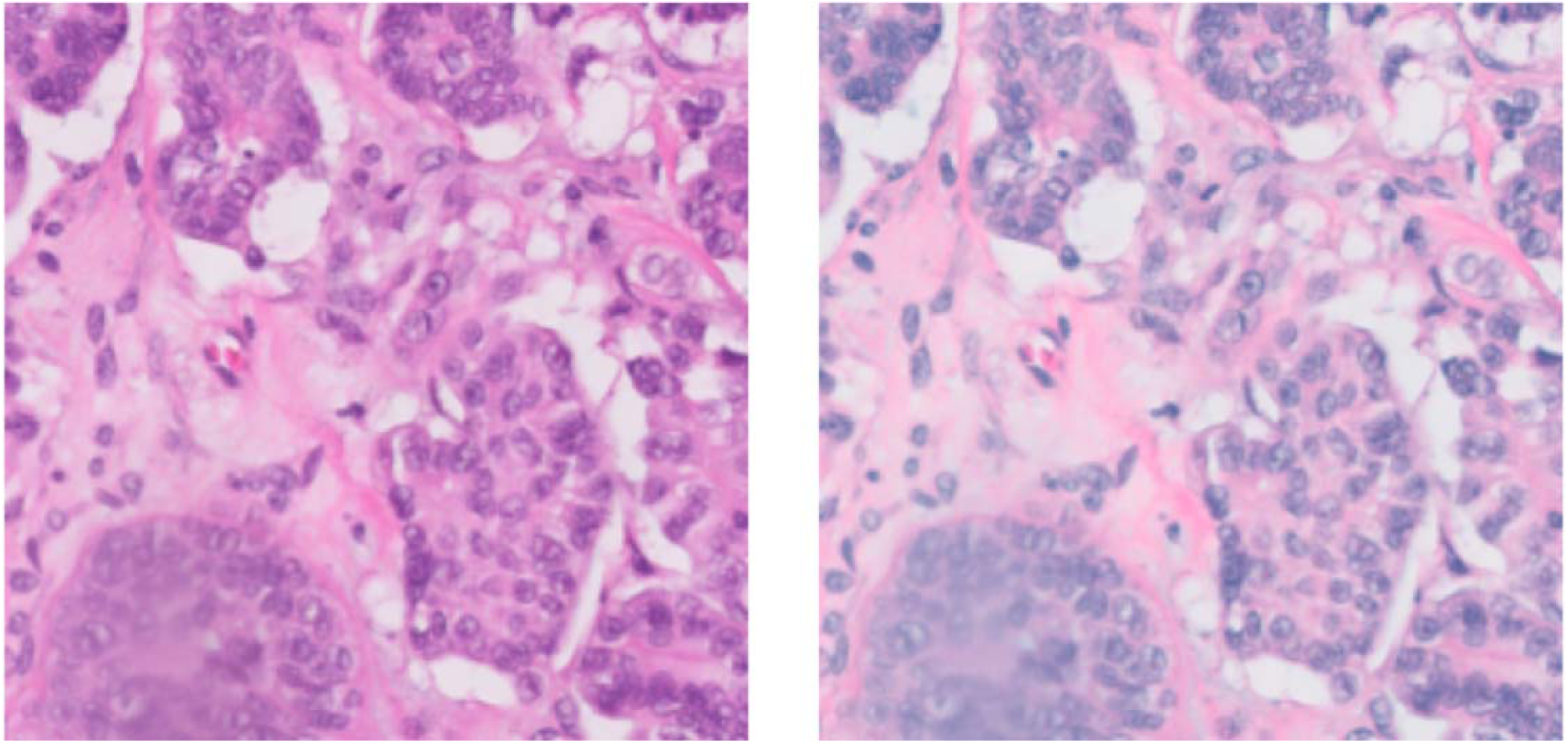
Representative tile before (left) and after (right) Macenko stain normalisation to a single reference stain matrix.

### 3.4 Models and Attention Mechanisms

To determine which attention design most benefits the HGSC-LGSC distinction, seven models were evaluated under an identical training and evaluation protocol. Five share one from-scratch four-block backbone and differ only in the attention block, forming a ladder from a no-attention baseline through Squeeze-and-Excitation, channel-only CBAM and full CBAM, with a lightweight CBAM variant isolating network size. The no-attention and channel-only models serve as ablation controls, the first isolating whether attention helps at all and the second whether the spatial branch adds anything beyond the channel descriptor. Two transfer-learning references complete the set, namely an ImageNet-pretrained MobileNet with a differential-attention module, and the Phikon pathology foundation model used as a frozen feature extractor with gated attention-based multiple-instance aggregation. All models were implemented in TensorFlow and Keras, trained with the same class-weighted binary cross-entropy loss, with the attention modules written as interchangeable layers so that a single block could be substituted without any other change.(30)(31)

#### 3.4.1 Attention modules

Three attention mechanisms were compared. The Convolutional Block Attention Module (CBAM) applies channel attention followed by spatial attention to recalibrate an intermediate feature map. Channel attention aggregates the map by global average and max pooling, passes both descriptors through a shared two-layer perceptron with reduction ratio r, sums them, and applies a sigmoid to obtain a per-channel weight, which rescales the feature map:(29)

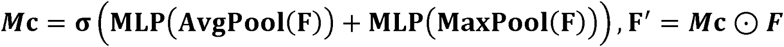

The channel-refined map is then reduced along the channel axis by average and max pooling, the two results are concatenated, and a 7×7 convolution with sigmoid activation produces a spatial weight map that rescales the map a second time:

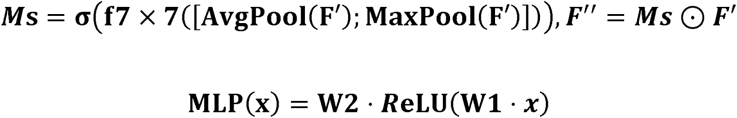

The Squeeze-and-Excitation (SE) block is a channel-only mechanism that aggregates the feature map by global average pooling and learns per-channel weights through a two-layer perceptron with a sigmoid gate, applying no spatial reweighting. Because SE is, in effect, the channel half of CBAM without its spatial branch, comparing the two under an otherwise identical backbone directly isolates the contribution of spatial attention. The differential-attention module takes a different approach, projecting the feature map into query, key, and value representations, computing dot-product attention, and applying a binary mask that retains only attention responses above their mean, which suppresses low-importance signal more sharply than the smooth sigmoid gating of CBAM and SE.

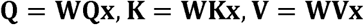

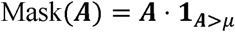

#### 3.4.2 Architectures

The deep CBAM and SE models share an identical four-block backbone. Each block applies a 3×3 convolution, batch normalisation, and a ReLU activation, followed by 2×2 max pooling and an attention module, with the number of filters doubling across blocks (32, 64, 128, and 256). The final feature map is reduced by global average pooling and passed through a 128-unit dense layer, dropout, and a single sigmoid output. Holding this backbone fixed, the three models differ only in their attention block. the deep CBAM model uses CBAM in each block, he lightweight CBAM model is a compact network of three convolutional blocks (8, 16, and 32 filters, reduction ratio 2) without the intermediate dense layer, totalling 7,955 parameters, with its configuration (dropout 0.3, learning rate 3×10⁻⁴) selected by a single-fold Hyperband search over filter counts, reduction ratio, dropout, and learning rate, to test how far performance can be preserved under a strict parameter budget; and the SE model is identical to the deep CBAM model except that CBAM is replaced by SE.

The pretrained model addresses a separate question, whether a pretrained backbone offers an advantage over the custom networks. It uses a MobileNet backbone pretrained on ImageNet and partially fine-tuned, with the differential-attention module applied over its final feature map before global average pooling, a dense layer, and a sigmoid output. Because MobileNet requires its own input scaling, its preprocessing differed accordingly, and its augmentation was limited to flips and ninety-degree rotations; all other elements of the protocol, including the data splits, loss, and slide-level evaluation, were held constant.

#### 3.4.3 Probability calibration

Neural network outputs are often poorly calibrated, meaning that predicted probabilities do not correspond to observed frequencies. To address this without altering the ranking of predictions, temperature scaling was applied as a post-hoc step to each trained model, dividing the logit of the slide-level probability by a single learned scalar T before the sigmoid:(35)

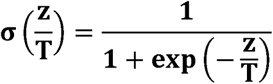

The temperature was fitted by cross-fitting across folds, estimating T for each fold from the out-of-fold predictions of the other four folds and applying it to that fold, so that no slide was calibrated with a temperature fitted on itself. Calibration was assessed by the expected calibration error (ECE) over ten equal-width probability bins, with a 95 percent bootstrap interval for its reduction:

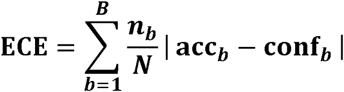

where n_b is the number of slides in bin b, and acc_b and conf_b are the observed LGSC frequency and mean predicted probability in that bin.

**Fig 3.4.**
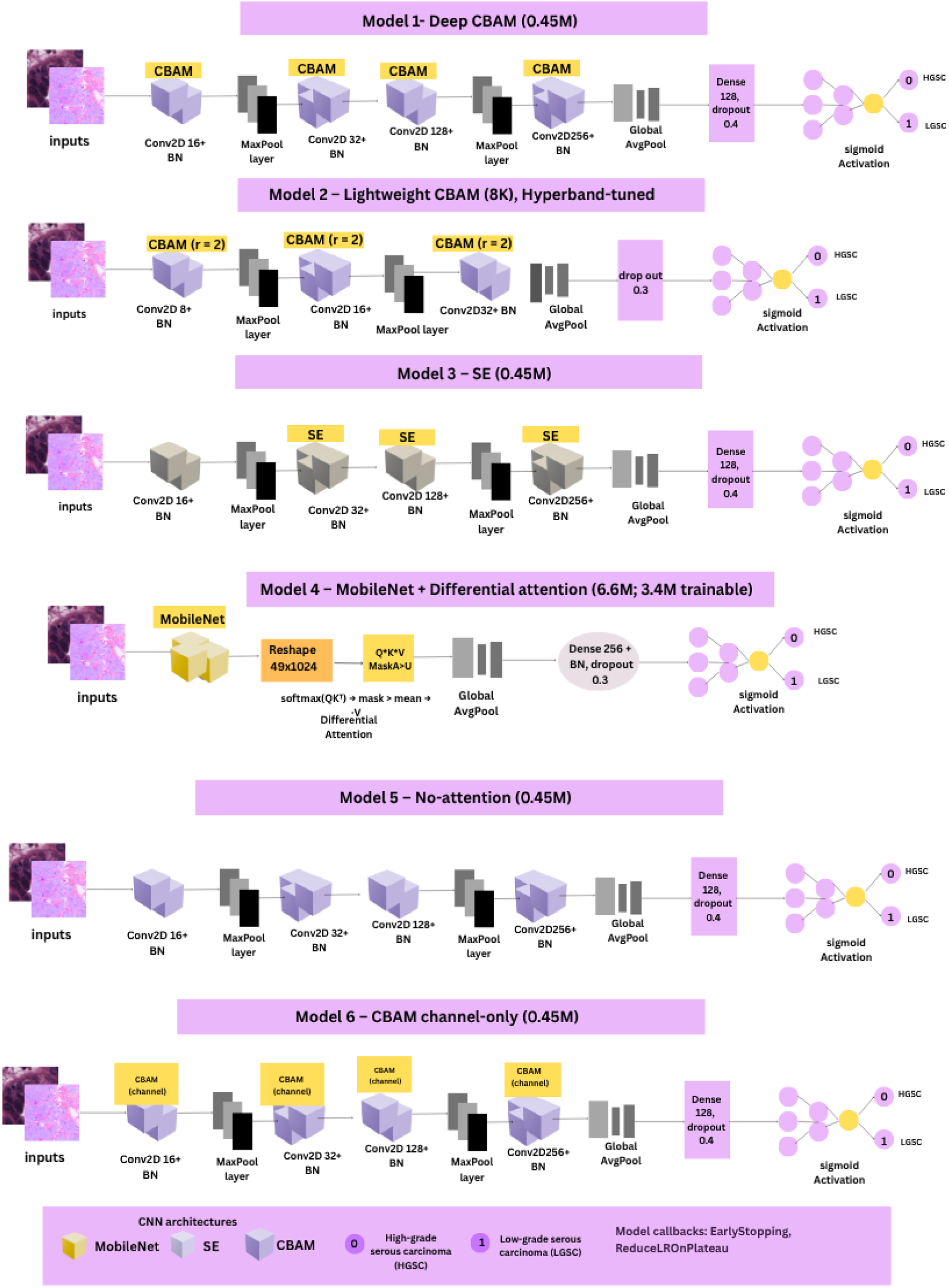
Architectures of the models. The five from-scratch models share one four-block backbone and differ only in the attention block (none, SE, channel-only CBAM, full CBAM); a lightweight three-block CBAM variant isolates network size. Model 4 applies differential attention to the final feature map of an ImageNet-pretrained MobileNet.

### 3.5 Training and Threshold Selection

All models were trained with a class-weighted binary cross-entropy loss, weighting each class inversely to its frequency so that the rare LGSC class is not overwhelmed during optimisation:

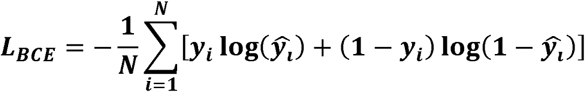

Optimisation used Adam (initial learning rate 1e-4, batch size 16) for up to 30 epochs, with early stopping on validation AUC, learning-rate reduction on a validation-loss plateau, and best-weight restoration; early stopping consistently halted training before the epoch limit. The lightweight model additionally underwent a single-fold Hyperband search, whose best configuration was applied across all folds. Tile probabilities were averaged to a slide-level prediction, and a single decision threshold was selected on the pooled out-of-fold predictions using Youden’s J statistic, which maximises sensitivity plus specificity; pooling the predictions before selecting the threshold avoids the instability of the few LGSC slides in any single fold.(34)

### 3.6 Evaluation Metrics and Statistical Analysis

Performance was summarised primarily by the area under the ROC curve (AUC), a threshold-independent measure of ranking ability that aligns with the DeLong test used for model comparison. The Matthews correlation coefficient (MCC), balanced accuracy, and the area under the precision-recall curve were reported as secondary, rare-class-sensitive measures. MCC uses all four confusion-matrix cells and remains informative under class imbalance, scoring near zero for a model that merely predicts the majority class:(33)

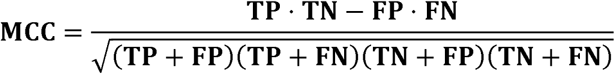

Per-class recall (sensitivity) was reported separately for each grade, defined as

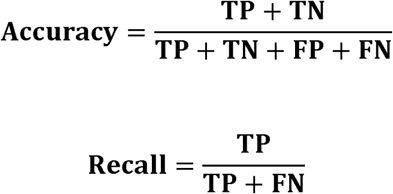

It is, however, threshold-dependent, and with only 42 LGSC slides the choice of operating point carries appreciable uncertainty; MCC is therefore reported alongside AUC rather than as the headline metric, since operating-point differences on this small sample can exceed genuine differences in ranking. AUC and MCC were reported with 95 percent confidence intervals from 2,000 bootstrap replicates resampled over slides, not tiles, since the slide is the independent unit of observation.

Model comparisons used two complementary tests, addressing the two distinct senses in which one model may exceed another. DeLong’s test compared AUCs, accounting for the correlation induced by evaluating both models on the same slides, and thus tested whether a difference in ranking is real:

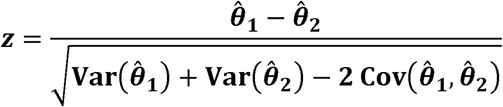

McNemar’s test compared slide-level decisions at each model’s threshold, examining only the slides on which the models disagree and testing whether that disagreement favours one model systematically:

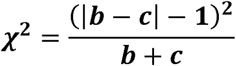

Reporting both distinguishes a genuine gain in ranking from a genuine gain in decisions, which need not coincide. Significance was set at p < 0.05, and as five pairwise comparisons were performed without correction for multiple testing, the resulting p-values are interpreted as exploratory. Because the LGSC count is small, these tests are conservatively powered, so a non-significant result indicates an absence of detectable difference rather than equivalence.

## 4. EXPERIMENTS AND RESULTS

Each model was trained and evaluated under the single leakage-controlled protocol described in Section 3: five-fold slide-grouped cross-validation on a common stain-normalised tile set, with patch predictions pooled to the slide level and decision thresholds selected within folds on validation slides. Every model was trained with three seeds, and all metrics are reported as mean and standard deviation across seeds, with pooled out-of-fold predictions providing one decision for all 259 slides. The from-scratch models share one four-block backbone and differ only in the attention block, forming a ladder from a no-attention baseline through Squeeze-and-Excitation, channel-only CBAM and full CBAM. A lightweight CBAM variant tests network size, and the pretrained MobileNet and the Phikon foundation model with attention-MIL provide transfer-learning references. Table 2 reports performance; Table 3 reports pairwise comparisons.(36)

**Table 2.**
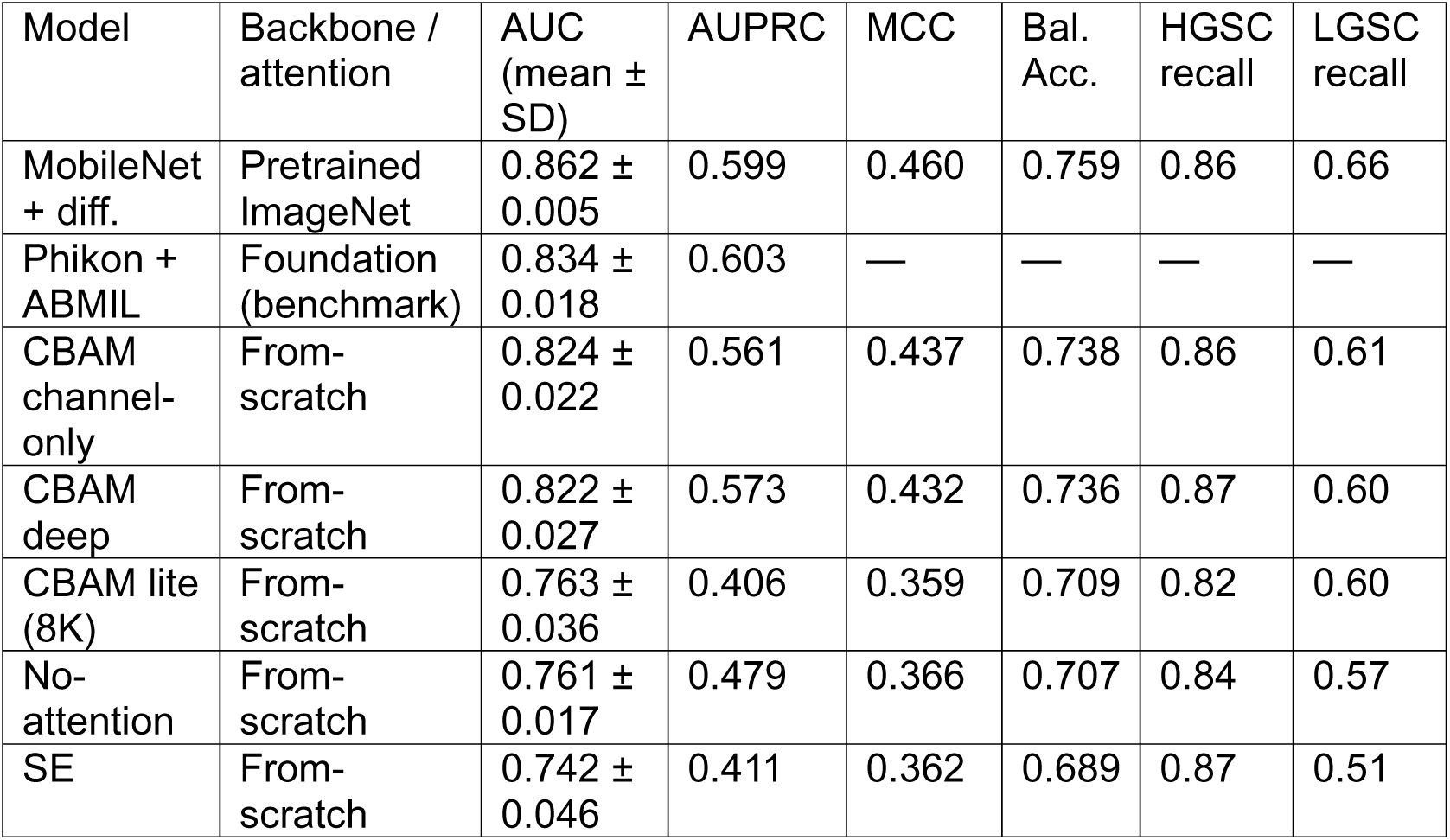
Slide-level performance on the binary HGSC versus LGSC task. Seven models were evaluated under one leakage-controlled protocol, with five-fold slide-grouped cross-validation, three training seeds, and the decision threshold selected within folds on validation slides. AUC and AUPRC are threshold-independent; MCC, balanced accuracy and per-class recall use the in-fold threshold. Values are mean ± SD over seeds. Colour-only baseline AUC 0.664 ± 0.082.

**Table 3.**
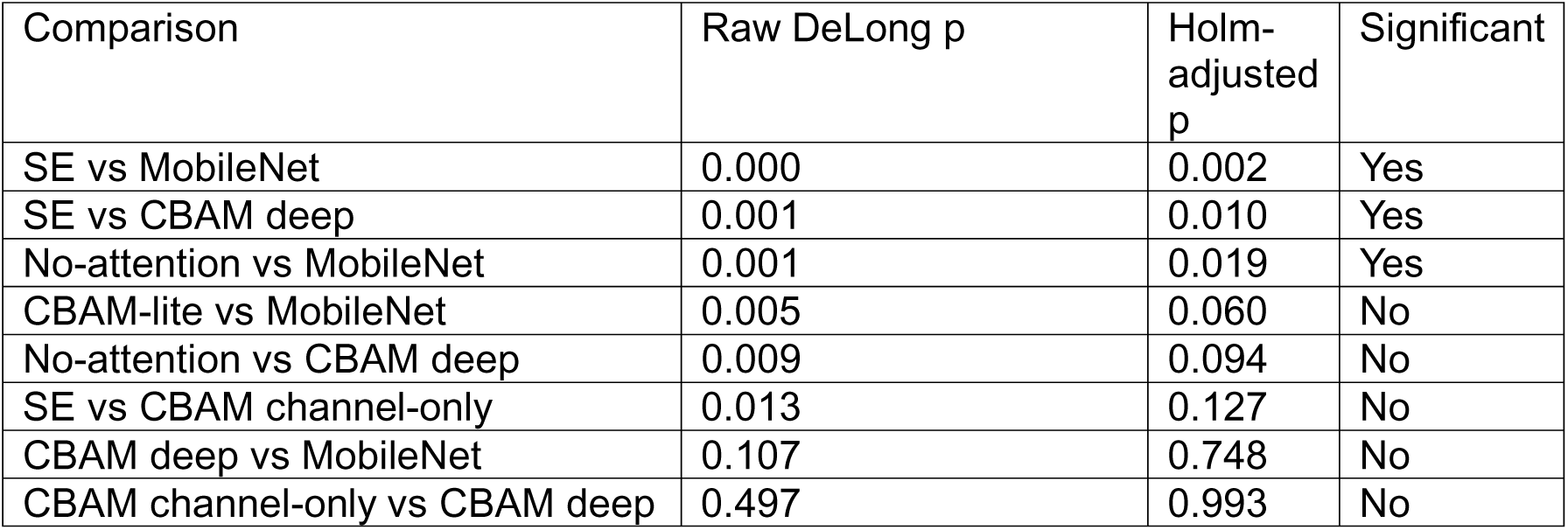
Pairwise AUC comparisons (DeLong), Holm-adjusted across the family. Only three survive at p < 0.05.

Temperature scaling was applied as a post-hoc calibration step and reduced the expected calibration error of the models while leaving AUC essentially unchanged. All fitted temperatures were below one, and some residual miscalibration remained after scaling, with the models tending to overestimate the probability of the rare low-grade class.(37)

The pretrained MobileNet reached the highest AUC, 0.862 ± 0.005, and was the most stable of all models, with a seed-to-seed standard deviation several times smaller than that of the from-scratch networks. It significantly outranked the Squeeze-and-Excitation model (DeLong, Holm p = 0.002) and the no-attention baseline (p = 0.019). Among the from-scratch models, full CBAM also outranked Squeeze-and-Excitation (p = 0.010), the only within-family comparison to survive correction; the pretrained model and full CBAM were not distinguishable from each other (p = 0.75). Adding the spatial-attention branch to channel attention changed AUC from 0.824 to 0.822 (p = 0.50), so the spatial branch carried no measurable benefit, and the gain over Squeeze-and-Excitation coincides with the avg-plus-max channel descriptor rather than the spatial branch. The lightweight model reached 0.763, close to the no-attention baseline and well below the deep CBAM networks, so the parity between a compact and a deep model reported under single-seed evaluation did not hold once training variance was included. The Phikon foundation model with attention-MIL reached 0.834 ± 0.018, below the pretrained MobileNet, so a pathology foundation model offered no advantage over ImageNet transfer on this binary task. A colour-only classifier, using only the RGB histogram of each tile, reached a slide-level AUC of 0.664 ± 0.082; the trained models exceed this but part of the separability is attributable to stain and colour rather than morphology. A per-branch analysis of mask-confirmed versus tissue-detected tiles was not conducted.

**Fig 4.1.**
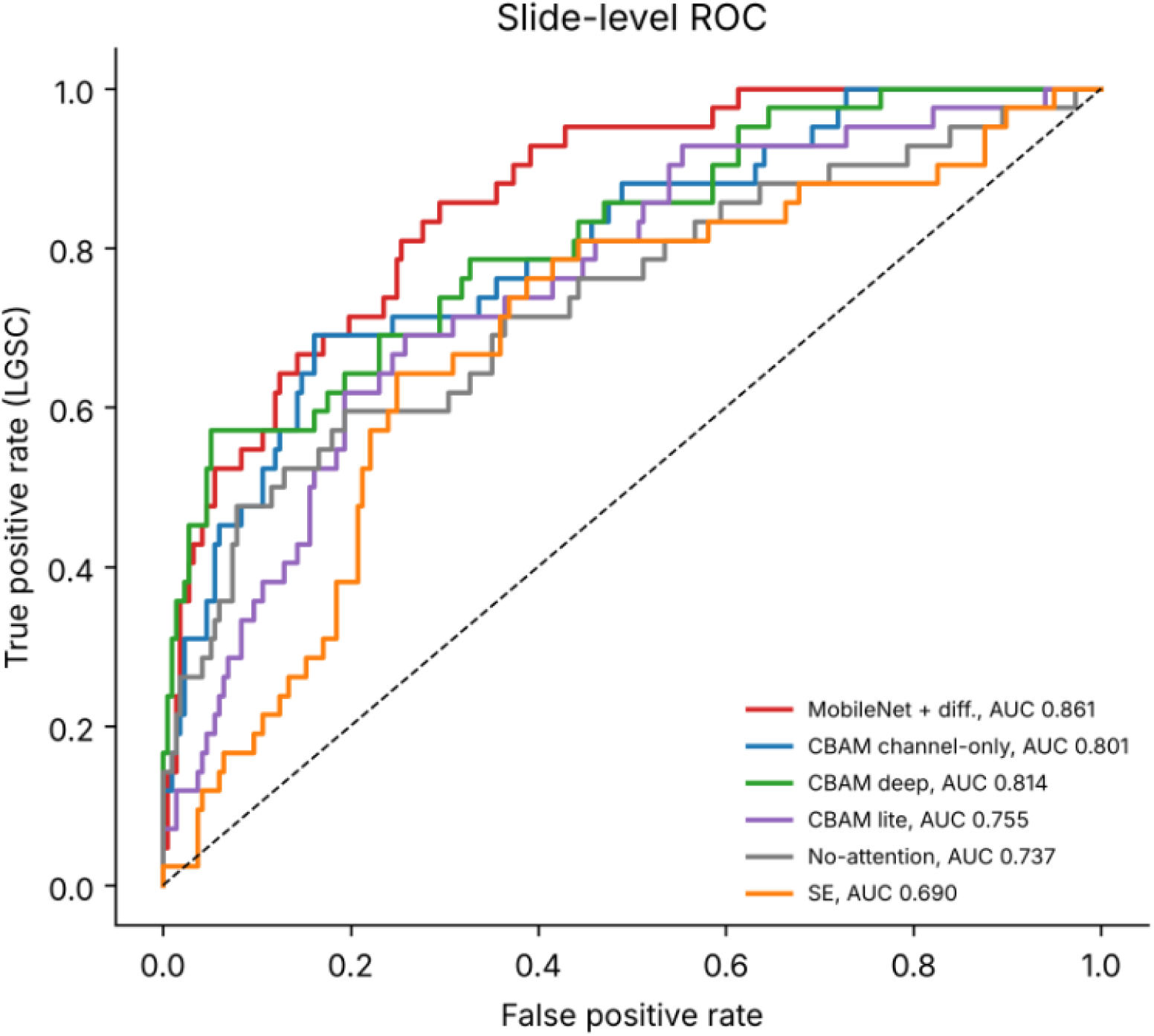
Slide-level receiver operating characteristic curves for the models, pooled across six cross-validation folds (n = 259 slides).

## 5. DISCUSSION

The seven models were designed to answer, under a common protocol, which attention mechanism and which feature source aid the high-versus low-grade serous distinction. Read together, they give a consistent account in which the source and reliability of the features matter more than the attention design or the network size. The clearest result is that transfer learning helped and was stable. The pretrained MobileNet reached the highest AUC and the smallest seed-to-seed variance, and it significantly outranked both the no-attention baseline and the Squeeze-and-Excitation model. For a cohort of 259 slides, which is too small to train good low- and mid-level filters from scratch, a backbone that already supplies descriptors of edges, textures and nuclear boundaries leaves only the task-specific head to be learned, and this shows in both the point estimate and the stability. The pretrained model did not, however, significantly exceed the best from-scratch network, so the honest statement is that transfer learning provided a reliable rather than a decisively larger improvement.

Within the from-scratch family, the one comparison that survived correction was full CBAM over Squeeze-and-Excitation. Because the two models share a backbone and differ in both the channel descriptor and the spatial branch, this gain cannot be attributed to a single component. The ladder narrows this further, since adding the spatial branch to channel attention produced no detectable change while the channel-only CBAM already matched the full model, which points to the avg-plus-max channel descriptor rather than the spatial branch as the active ingredient. With 42 low-grade slides the study is underpowered to confirm this at the level of the single component, and the intermediate comparisons did not reach significance. The spatial-attention branch, which recent work has treated as the main contribution of CBAM over Squeeze-and-Excitation, did not help here.

Network size was not a factor. The lightweight model, at roughly 8000 parameters, fell to the level of the no-attention baseline and below the deep CBAM networks once training variance was included, so the apparent parity between a compact and a deep model seen under a single seed did not survive repeated runs. This is a caution against reading a single-seed result on a small rare class as evidence of a capacity effect.(25)

The foundation-model benchmark is the most informative negative result. A pathology foundation model, trained with self-supervision on far larger cohorts than ImageNet, reached 0.834 and did not exceed the pretrained MobileNet. On the five-class subtyping task the literature reports that the choice of feature extractor dominates performance; on this binary serous grade distinction, under leakage-controlled evaluation on a small cohort, that advantage did not appear. The ceiling of roughly 0.83 to 0.86 across the strongest models, rather than the particular backbone, is the recurring feature of the results, and it is consistent with the difficulty of the distinction and the scarcity of the rare class.

A controlled comparison shows which design performs better, but not whether a model reaches its decisions for the right reasons. A colour-only baseline reached a slide-level AUC of 0.66, so part of the separability is carried by stain and colour rather than morphology, even after Macenko normalisation. The trained models exceed this baseline and therefore use more than colour, but the gap between 0.66 and roughly 0.85 bounds the morphological component of their performance and is a reminder that multi-centre colour variation is a plausible partial shortcut on this cohort. The per-class recall confirms that the pretrained model’s advantage is specific to the rare class. It reached the most balanced recall across grades, 0.86 for high-grade and 0.66 for low-grade carcinoma, while the Squeeze-and-Excitation model had the weakest low-grade recall at 0.51. Against a low-grade prevalence of 0.16, the pretrained model and the foundation model reached the highest average precision, around 0.60, which shows their advantage lies in the rare class rather than the majority. With only 42 low-grade slides even a handful of ambiguous cases can shift a threshold-dependent metric, which accounts for the wide intervals and the fact that most comparisons did not reach significance.

These architectural findings carry weight only because they were obtained under a protocol that allows the seven models to be compared on equal terms, and the case for that protocol becomes clear when the existing results on UBC-OCEAN are placed side by side. The dataset was assembled from more than twenty centres with deliberately heterogeneous tissue processing and staining, and its organisers evaluated the challenge on a held-out test set kept separate from algorithm development, explicitly to mitigate the risk of information leakage. The dataset was, in other words, designed with leakage in mind, and the results reported on it vary with how closely later studies have followed that design.(13)

**Fig 4.2.**
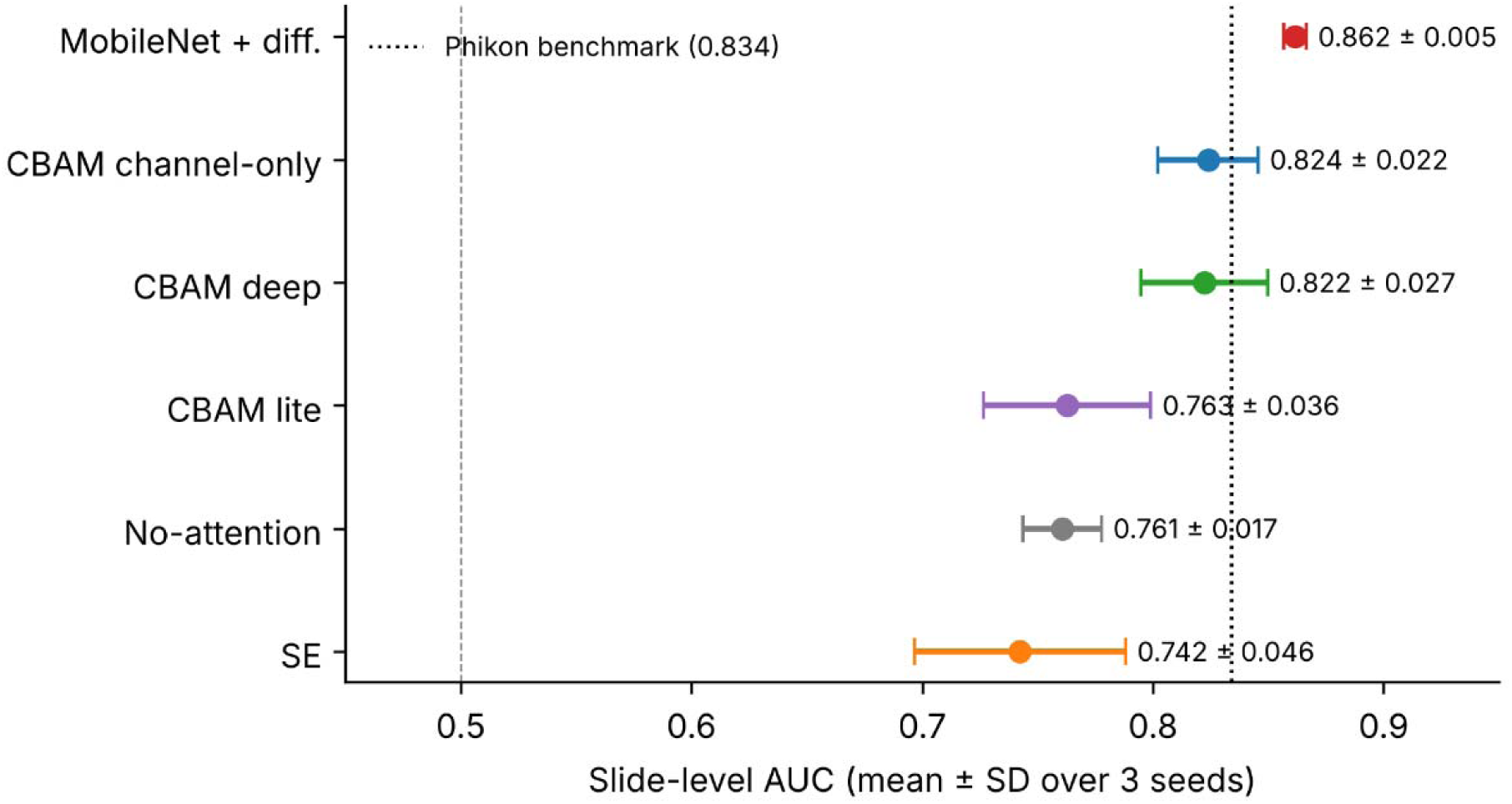
Slide-level AUC with 95% bootstrap confidence intervals, resampled over slides (2,000 replicates).

**Fig 4.3.**
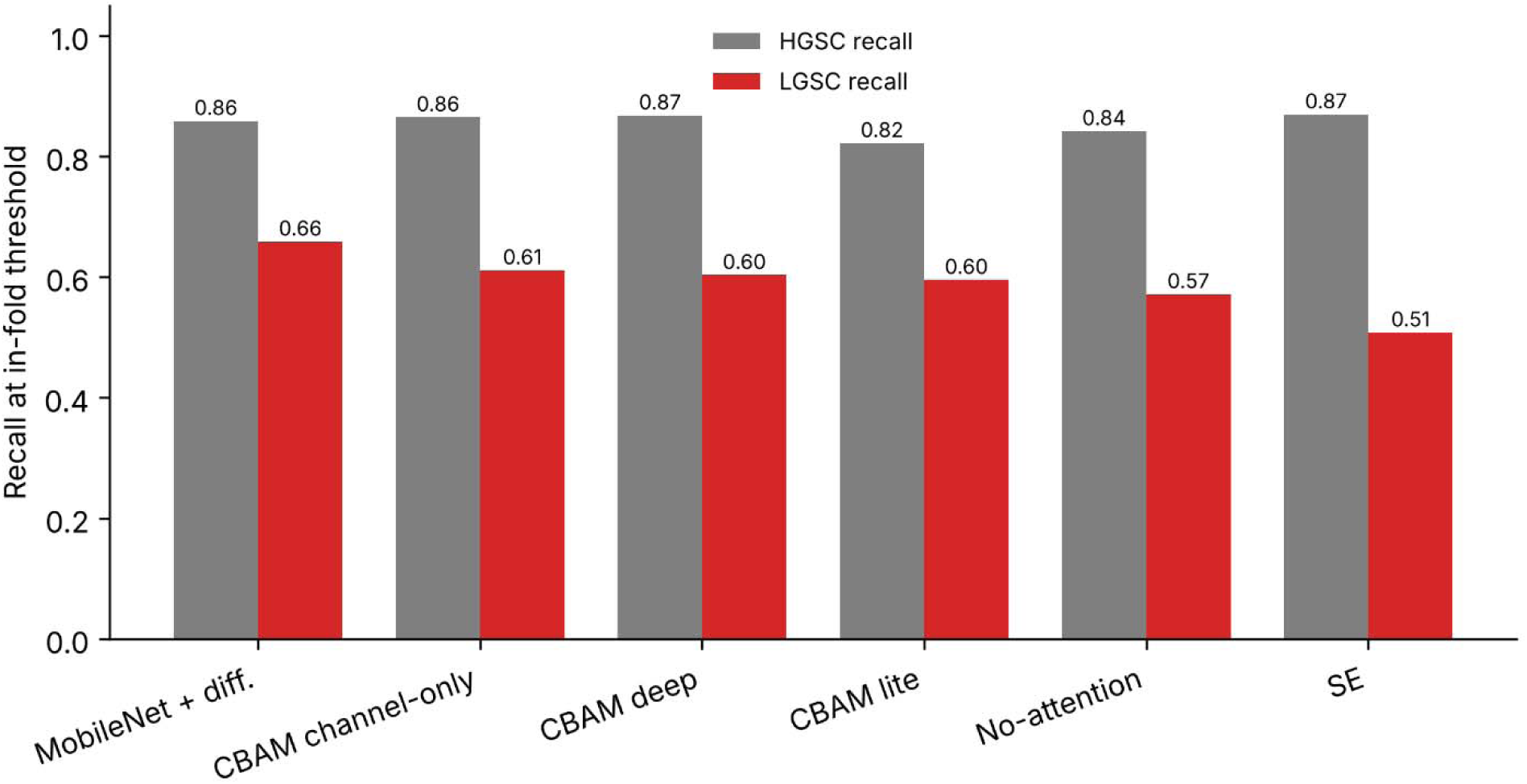
Per-class recall for high-grade (HGSC) and low-grade (LGSC) serous carcinoma for each model, at the Youden-optimal threshold.

Arranged by the strictness of their evaluation, the published results show a consistent pattern (Table 4). On the organisers’ own held-out multi-centre test set, the top-performing solutions to the five-class challenge reached balanced accuracies of only 58 to 66 percent (13). Breen et al., training on 1,864 whole-slide images from an independent centre and validating externally on 513 OCEAN slides, reported a best balanced accuracy of about 0.83 with a self-supervised foundation-model extractor, falling to roughly 0.80 on the external OCEAN cohort, and noted that the dataset’s highly heterogeneous preparation contributed to the difficulty (10). The same evaluation found performance to depend strongly on the feature extractor, with the strongest extractors well ahead of ImageNet-pretrained baselines, and summarised earlier cross-validated studies on comparably sized ovarian cohorts reporting balanced accuracies in the region of 0.73 to 0.83 (10). At the other end, results obtained under within-dataset splits drawn at the level of tiles or images are considerably higher. TopOC, trained and tested on 34,000 tiles derived from 975 whole-slide and tissue-microarray images under a predefined tile-level split, reported 66 percent balanced accuracy and compared it with the challenge-winning 66 percent, a comparison between a tile-level internal split and a held-out external evaluation (27). Behera et al. reported complete accuracy under a single image-level 80-to-20 split without cross-validation, while their own per-class results show a high-grade serous sensitivity of 42.9 percent (26). Several of the highest accuracies cited for ovarian subtyping were not obtained on this dataset at all. OvCan-FIND’s 99.74 percent, for example, comes from 5,942 images of a single-centre cohort (14). These figures are five-class balanced accuracies and are not directly comparable with the binary AUC reported here; the point is the gradient itself. The more closely an evaluation separates training and test data by case and by centre, the lower the reported performance, which indicates that the difficulty of the task has often been understated.(27)(10)(10)(10)(10)(15)

**Table 4.** Evaluation protocols of representative studies, ordered from strictest to least strict. Reported values are five-class balanced accuracy unless stated, and are not directly comparable with the binary AUC of the present study.

| Study | Cohort used | Unit of split | Validation | Reported result |
|---|---|---|---|---|
| OCEAN challenge, top solutions (13) | Organisers' held-out multi-centre test set | Case / centre | External, held out by organisers | BAcc 58–66% |
| Breen et al. (10) | Trained on 1,864 Leeds WSIs; tested on 513 OCEAN WSIs | Slide | External | BAcc ~0.80 on OCEAN (best extractor ~0.83) |
| Cross-validated MIL (summarised in 10) | 538 OCEAN WSIs | Slide | 10-fold CV + external (TCGA) | BAcc ~0.73–0.83 |
| Feature-extractor comparison (summarised in 10) | 510 OCEAN WSIs | Slide | Internal | BAcc varies strongly with extractor |
| (38) | TopOC (27): 34,000 tiles from 975 WSIs/TMAs | Tile | Predefined internal split | BAcc 66% |
| (27) | Behera et al. (26): 725 images (5-class) | Image | Single 80/20 split, no CV | Accuracy 100% (HGSC sensitivity 42.9%) |
| Present study | 259 WSIs, 20x only, binary HGSC vs LGSC | Slide | 5-fold slide-grouped CV | AUC 0.74–0.86 (3 seeds, mean±SD) |

Two further features of the literature prevent these results from being compared at all. First, there is no canonical form of the benchmark. The same dataset appears as 505, 510, 513, 538, or 975 images, with whole-slide images at 20x and tissue microarrays at 40x sometimes pooled and sometimes separated, and with tiles, images, or slides used interchangeably as the unit of analysis. Each study therefore evaluates a differently derived subset under a differently constructed protocol, so two reported numbers need not measure the same quantity. Second, some properties of the data remain undocumented, including how ground-truth labels were determined and which tissue types were included, so that differences between studies cannot always be traced to their source. The finding that the choice of feature extractor alone can move balanced accuracy across nearly the entire achievable range makes the point directly, since when a single uncontrolled component produces a swing of that size, a bare accuracy figure says little about a method unless everything else is held fixed.(10)

The design of this study was shaped throughout by a single goal, to make each reported difference interpretable rather than merely favourable. Several recurring weaknesses in the existing literature motivated the specific choices. Pooling whole-slide images with tissue microarrays mixes two magnifications and lets scale, rather than morphology, drive part of the separation, so the cohort was restricted to 20x whole-slide images; the trade-off is that resizing tiles to 224 pixels yields an effective resolution of about 1.14 micrometres per pixel, coarser than the acquisition itself. Tile- and image-level partitioning allows patches from one slide to fall on both sides of the split, so a model can recognise slide identity instead of disease, and assigning every tile of a slide to a single partition closes that route. Averaging the rare grade into a five-class score lets a weak class hide behind a strong aggregate, so isolating the binary grade distinction exposes low-grade performance directly, where it is hardest and most clinically consequential. Centre-specific staining is a plausible shortcut on a multi-centre cohort, so normalising every tile against one fixed reference reduces that variation without carrying any label information. Finally, single point estimates on 42 low-grade slides are unstable, so reporting standard deviations across seeds, bootstrap confidence intervals over slides, and Holm-corrected tests replaces them with bounded, corrected quantities. Together these choices are what give the comparison its force, since backbone, loss, data and evaluation are held identical and only the attention block or the backbone varies, so each difference in performance can be attributed to one design choice, which is the defining property of a controlled ablation. The resulting AUCs of 0.74 to 0.86 sit well below the near-perfect figures reported under tile- and image-level protocols, which is the expected consequence of removing those shortcuts rather than a sign of a weaker method. The one gap these controls do not address is external generalisation, since the results come from within-cohort cross-validation rather than an independent test set, and external validation is therefore the natural next step.

These findings should also be read against how the distinction is made in practice. Pathologists separate the two grades using the two-tier system, which rests on nuclear atypia and mitotic activity assessed at high magnification. At the effective resolution used here, individual mitoses and nucleoli are at or below the limit of reliable resolution, so the models most plausibly draw on architectural pattern, nuclear crowding, and gross variation in nuclear size rather than on the criteria a pathologist counts, and they therefore approximate the grading decision rather than reproduce it. Such a system is best suited to an assistive role, flagging slides whose features suggest low-grade morphology for ancillary immunohistochemistry or specialist review where gynaecological pathology expertise is limited. The balanced recall of the pretrained model matters most for this role, since a triage tool must recover the rare class without excessive false alarms on the common one. Temperature scaling improved calibration, but every model continued to overestimate the probability of low-grade carcinoma, a bias that a single temperature cannot correct and that a method with an intercept term, such as Platt scaling, could address. The competitiveness of the lightweight network is relevant here too, since an 8,000-parameter model can run on modest hardware where larger networks are impractical.

Several limitations bound these conclusions, and they also frame how the moderate values reported here should be read. The performance of all models is consistent with other case-level evaluations on this dataset, and the much wider range of figures in the literature appears to reflect differences in evaluation design as much as in method. The cohort is the principal constraint, with only 42 low-grade slides, which widens every confidence interval and leaves most comparisons short of significance after correction. All results come from within-cohort cross-validation on a single dataset, so external validation on independent cohorts with adequate low-grade representation remains necessary, and the scarcity of public low-grade slides currently limits it. The pretrained model also differed from the from-scratch models in both backbone and attention type, so its advantage cannot be cleanly separated between the two, and the coarse effective resolution of the resized tiles further restricts access to the nuclear detail on which grading depends. A further limit concerns interpretability, since the present analysis establishes which design ranks higher but not whether a model attends to tumour tissue, and a direct evaluation of the attention modules against expert masks, using localisation metrics such as the pointing game and per-pixel saliency agreement, is the natural next step, with the colour-only control already bounding how much of the separability is non-morphological. Each of these points suggests a direct extension, namely external testing on multi-centre cohorts, higher-magnification inputs that resolve nuclear and mitotic detail, pathology-specific foundation-model features with attention-based aggregation, and finer-grained attribution applied to the attention modules. Taken together, the study shows that under leakage-controlled evaluation the high-versus low-grade distinction remains genuinely difficult, that transfer learning and the avoidance of a weak channel-only configuration contribute measurably while spatial attention and network size do not, and that these conclusions become clear only when every model is measured against the same fixed standard.

## 6. CONCLUSION

Automated grading of serous ovarian carcinoma matters because the high-versus low-grade distinction directly shapes treatment, yet it is the boundary that both pathologists and algorithms find hardest. This work compared attention and architecture choices one at a time under a common, leakage-controlled standard, across three seeds and with multiple-comparison correction, and showed that the distinction is more difficult than much of the literature suggests. Under these conditions the source and reliability of the features mattered more than the attention design or the network size. Transfer learning and the avoidance of a weak channel-only configuration produced the clearest gains, while the spatial attention branch, a larger network and a pathology-specific foundation model produced no detectable improvement. These conclusions rest on a single cohort of 42 low-grade slides, which leaves most finer comparisons underpowered. Future work should pursue external validation on cohorts with greater low-grade representation, higher-magnification inputs capable of resolving mitotic detail, direct evaluation of the attention modules against expert masks, and a shared evaluation protocol for UBC-OCEAN that allows methods to be compared directly.

## Code and Data Availability

The primary dataset used in this study is openly accessible via the UBC Ovarian Cancer Subtype Classification and Outlier Detection (UBC-OCEAN) competition, hosted by the Artificial Intelligence in Medicine (AIM) Lab at the University of British Columbia: https://kaggle.com/competitions/UBC-OCEAN. No additional datasets were generated during this study.

The complete source code, model implementations, attention module configurations, and experimental scripts are publicly available at: https://github.com/AshsmashR/CNN-Attention-Ovarianhistopath. The repository includes all training and evaluation pipelines necessary to reproduce the results reported in this article.https://github.com/AshsmashR/CNN-Attention-Ovarianhistopath

## Data Citation

Asadi-Aghbolaghi, M., Farahani, H., Zhang, A., Akbari, A., Kim, S., Chow, A., Dane, S., OCEAN Challenge Consortium, OTTA Consortium, Huntsman, D. G., Gilks, C. B., Ramus, S., Köbel, M., Karnezis, A. N., & Bashashati, A. (2024). Machine learning-driven histotype diagnosis of ovarian carcinoma: Insights from the OCEAN AI Challenge. *medRxiv*. https://doi.org/10.1101/2024.04.19.24306099

UBC Ovarian Cancer Subtype Classification and Outlier Detection (UBC-OCEAN) [Dataset]. (2023). Kaggle. https://kaggle.com/competitions/UBC-OCEAN

## Acknowledgements

The authors thank Mr Sagar (MSc, DPSRU, New Delhi) for his assistance with data preprocessing, and Ms Aparna Singh for their critical reading and review of the manuscript.

## Author Information

Competing Interests: The authors declare no competing interests.

## Author Contributions

A.R.: Conceptualization, Methodology, Software, Formal analysis, Investigation, Data curation, Visualization, Writing – review & editing. S.M.: Validation, Writing – original draft, Writing – review & editing, Project administration. Both authors contributed equally and share co-first authorship. Both authors read and approved the final manuscript.

## Ethics Declarations

This article does not contain any studies with human participants or animals performed by any of the authors.

